# Integration of multi-omics data improves prediction of cervicovaginal microenvironment in cervical cancer

**DOI:** 10.1101/2020.08.27.20183426

**Authors:** Nicholas A. Bokulich, Paweł Łaniewski, Dana M. Chase, J. Gregory Caporaso, Melissa M. Herbst-Kralovetz

## Abstract

Emerging evidence suggests that a complex interplay between human papillomavirus (HPV), microbiota, and the cervicovaginal microenvironment contribute to HPV persistence and carcinogenesis. Integration of multiple omics datasets is predicted to provide unique insight into HPV infection and cervical cancer progression. Cervicovaginal specimens were collected from a cohort (n=100) of Arizonan women with cervical cancer, cervical dysplasia, as well as HPV-positive and HPV-negative controls. Microbiome, immunoproteome and metabolome analyses were performed using 16S rRNA gene sequencing, multiplex cytometric bead arrays, and liquid chromatography-mass spectrometry, respectively. Multi-omics integration methods, including neural networks (mmvec) and Random Forest supervised learning, were utilized to explore potential interactions and develop predictive models. Our integrated bioinformatic analyses revealed that cancer biomarker concentrations were reliably predicted by Random Forest regressors trained on microbiome and metabolome features, suggesting close correspondence between the vaginal microbiome, metabolome, and genital inflammation involved in cervical carcinogenesis. Furthermore, we show that features of the microbiome and host microenvironment, including metabolites, microbial taxa, and immune biomarkers are predictive of genital inflammation status, but only weakly to moderately predictive of cervical cancer state. Different feature classes were important for prediction of different phenotypes. Lipids (e.g. sphingolipids and long-chain unsaturated fatty acids) were strong predictors of genital inflammation, whereas predictions of vaginal microbiota and vaginal pH relied mostly on alterations in amino acid metabolism. Finally, we identified key immune biomarkers associated with the vaginal microbiota composition and vaginal pH (MIF and TNFα), as well as genital inflammation (IL-6, IL-10, leptin and VEGF). Integration of multiple different microbiome “omics” data types resulted in modest increases in classifier performance over classifiers trained on the best performing individual omics data type. However, since the most predictive features cannot be known *a priori*, a multi-omics approach can still yield insights that might not be possible with a single data type. Additionally, integrating multiple omics datasets provided insight into different features of the cervicovaginal microenvironment and host response. Multi-omics is therefore likely to remain essential for realizing the advances promised by microbiome research.

## Background

Despite the availability of preventive measures, such as routine human papillomavirus (HPV) vaccination and Pap smear screening, cervical cancer remains a major public health problem, particularly in low- and middle-income countries, with approximately 570,000 new cases and 311,000 deaths worldwide in 2018 (1). From an epidemiological standpoint, infection with high-risk HPV types is a well-established risk factor for cervical cancer (2). Genital HPV infection, although necessary, is not sufficient for development of precancerous cervical dysplasia and progression to cancer (3), suggesting that other factors in the local cervicovaginal microenvironment play a role during cervical carcinogenesis (4).

In the last two decades, the human microbiome (collectively the microbiota, or communities of microorganisms residing in and on the human body, and their theatre of activity (5) has emerged as a key regulator of mucosal homeostasis at various body sites, including the female reproductive tract (6). The cervix and vagina in the majority of healthy, reproductive-age women are colonized by one or few *Lactobacillus* species, primarily *L. crispatus, L. iners, L. gasseri*, or *L. jensenii* (7). These beneficial microorganisms produce lactic acid (lowering vaginal pH, typically below 4.5) and other antimicrobial metabolites, as well as block attachment of other bacteria to the genital epithelium through competitive exclusion mechanisms. In addition, *Lactobacillus* spp. stimulate the host to secrete physiological levels of cytokines, antimicrobial peptides and metabolites (8).

Collectively, multifaceted interactions between *Lactobacillus* and the host create a protective microenvironment against invading bacteria, fungi and viruses, including HPV (9). However, during dysbiosis (disruption of the local microbial ecosystem, such as during disease) protective *Lactobacillus* spp. are depleted and replaced by a diverse consortium of obligate and strict anaerobes, resulting in elevated vaginal pH (10). These changes are associated with increased risk for adverse gynecologic and reproductive outcomes, including sexually transmitted infection (STI) acquisition (11). Indeed, several clinical studies have demonstrated that HPV infection associates with substantial changes in the cervicovaginal microenvironment, including shifts in microbial (12-28), metabolic (29, 30) and immunoproteomic profiles (17, 18, 31, 32), as well as vaginal pH levels (17), which might drive HPV persistence and/or disease progression.

Multiple cross-sectional studies in various racial/ethnic cohorts consistently demonstrated that women infected with HPV exhibit more diverse vaginal microbiota and depleted levels of beneficial *Lactobacillus* spp. compared to HPV-negative women (12-15). Women with cervical dysplasia or cancer also commonly lacked *Lactobacillus* dominance in their vaginal microbiota (16-21). Furthermore, bacterial vaginosis (BV), which is a common vaginal disorder characterized by a dramatic shift in microbiota composition from *Lactobacillus* to anaerobes, has been linked to an increased risk of HPV acquisition and persistence (33-35). Limited longitudinal studies also demonstrated that *Lactobacillus-dominant* microbiota correlates with HPV clearance and regression of dysplasia, whereas depletion of *Lactobacillus* and presence of specific anaerobic bacteria is associated with HPV and disease persistence (22-25). Recent systematic reviews and meta-analyses of available studies supported a causal link between dysbiotic vaginal microbiota and cervical cancer through the impact of bacteria on HPV acquisition and persistence, as well as dysplasia development (26-28).

Metabolically, limited studies have reported that HPV infection and cervical dysplasia relate to depletion of amino acid, peptide, and nucleotide signatures in the cervicovaginal microenvironment (29, 30). Intriguingly, these metabolic alterations are also associated with depletion of *Lactobacillus* spp., connecting HPV infection to vaginal dysbiosis (29, 36). In contrast, cervical carcinoma profoundly perturbs lipid signatures, such as sphingomyelins (29), which are also biomarkers of chronic inflammation (37) and associated with genital inflammation (29).

In regard to host immune defenses, it is well documented that persistent HPV infection suppresses immune responses, which may contribute to progression of HPV-mediated neoplasm (38). Yet, the impact of the microbiome on host defenses across cervical carcinogenesis has not been comprehensively studied. Recent studies have revealed that dysbiotic *non-Lactobacillus* dominant microbiota are associated with elevated levels of pro-inflammatory cytokines, growth and angiogenesis factors, apoptosis-related proteins, and immune checkpoint proteins in the cervicovaginal fluids (17, 31, 32). Another cross-sectional study suggested a link between dysbiotic fusobacteria and immunosuppressive host responses (18). Taken together, these reports strongly implicate the complex interplay between HPV, microbiota, and host response mechanisms in the local microenvironment in the progression of (or protection from) neoplastic disease.

Here we present an integrated multi-omics analysis of clinical datasets including vaginal microbiome (17), vaginal pH (17), metabolome (29) and immunoproteome (17, 31, 32), which were previously generated using cervicovaginal specimens collected from a cohort (*n*=100) of women with cervical cancer, cervical dysplasia, as well as HPV-positive and HPV-negative controls, but which were previously analyzed independently. Cervical cancer disease phenotypes emerge from the interactions between multiple features, including microbial taxa, metabolic activity of microbes, host immune system activity, and the vaginal microenvironment. Hence, we hypothesized that applying newly developed multi-omics integration techniques, including microbe–metabolite vectors (mmvec (39)) neural networks and Random Forest supervised learning models to delineate relationships between microbial, metabolic, and proteomic signatures across a cervical carcinogenesis spectrum would allow us to learn more from these data than we could from any single data type in isolation. We present new predictive models of *Lactobacillus* dominance, vaginal pH, genital inflammation and cervical neoplastic disease, and discuss the relative contribution of different features and feature types to our top-performing models (**Figure 1**).

**Figure 1.**
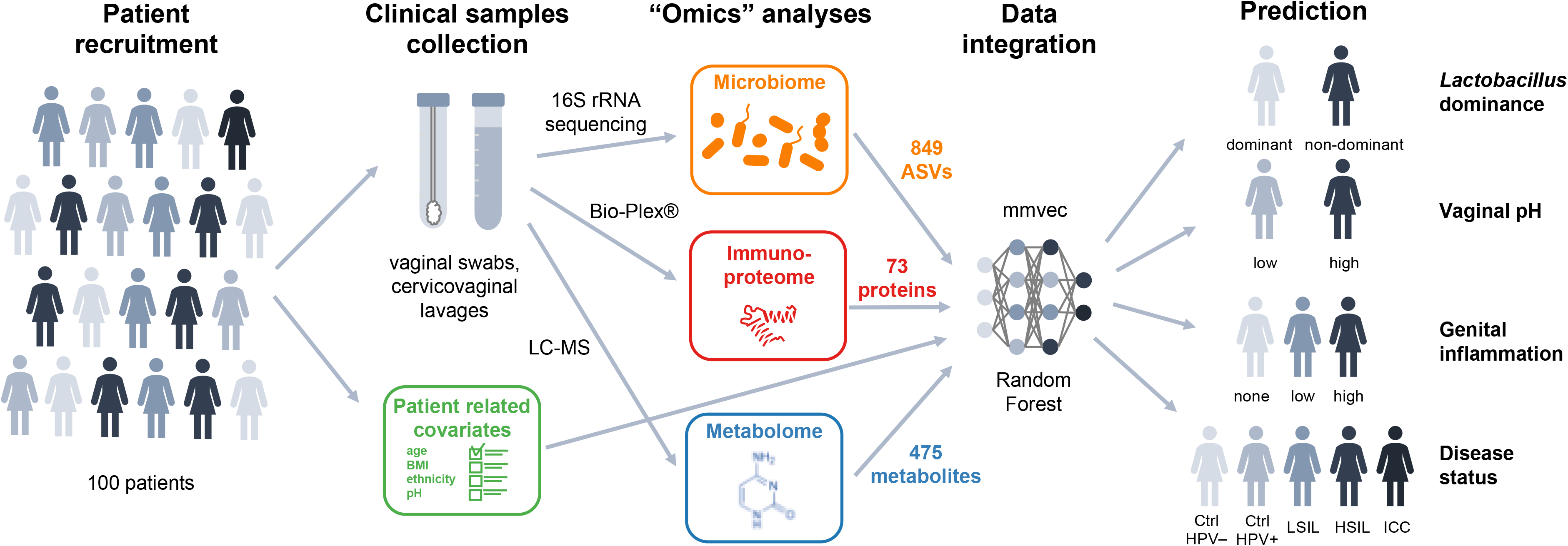
Schematic of a multi-omics approach to study the complex interplay between HPV, host and microbiota in women across cervical neoplasia. In this multicenter study *n*=100 women were enrolled with invasive cervical carcinoma (ICC), high- and low-grade squamous intraepithelial lesions (HSIL, LSIL), as well as, HPV-positive and healthy HPV-negative controls (Ctrl). Vaginal swabs and cervicovaginal lavages (CVL) were collected for vaginal pH, microbiome, metabolome and immunoproteome analyses. Patient-related metadata, including age, body mass index (BMI), ethnicity, were also collected through medical records and surveys. The vaginal microbiota compositions were determined by 16S rRNA gene sequencing (*n*=99) revealing 849 amplicon sequencing variants (ASVs). Cervicovaginal metabolic fingerprints were profiled by liquid chromatography-mass spectrometry (*n*=78) and identified 475 unique metabolites. Levels of immune mediators (*n*=100) and other cancer-related proteins (*n*=78) in CVL samples (73 targets) were evaluated using multiplex cytometric bead arrays. Principal component, hierarchical clustering, neural network (mmvec) and Random Forest analyses were utilized to explore associations among multi-omics data sets to predict *Lactobacillus* dominance (dominant vs. non-dominant), vaginal pH (low ≤5 vs. high >5), evidence of genital inflammation (high, low, none) and disease status (Ctrl HPV−, Ctrl HPV+, LSIL, HSIL, ICC).

## Methods

### Study population and clinical sample collection

One hundred premenopausal, non-pregnant women were recruited at three clinical sites located in Phoenix, AZ: St. Joseph’s Hospital and Medical Center, University of Arizona Cancer Center and Maricopa Integrated Health Systems. All participants provided informed written consent and all research and related activities involving human subjects were approved by the Institutional Review Boards at each participating site. The participants were grouped as follows: HPV-negative controls [Ctrl HPV- (*n*=20)], HPV-positive controls [Ctrl HPV+ (*n*=31)], low grade squamous intraepithelial lesions [LSIL (*n*=12)], high grade squamous intraepithelial lesions [HSIL (*n*=27)] and invasive cervical carcinoma [ICC (*n*=10)]. Classification of patients into the five groups and detailed exclusion criteria were described previously (Łaniewski et al., 2018). Cervicovaginal lavage (CVL) and vaginal swabs were collected by a physician and processed as described previously (17). Vaginal pH was measured using vaginal swabs, nitrazine paper and a pH scale ranging from 4.5 to 7.5 (17). Demographic data was collected from surveys and/or medical records.

### Immunoproteome analysis

Levels of 73 protein targets were determined in CVL samples using multiplex cytometric bead arrays or enzyme-linked immunosorbent assays and described previously (17, 31, 32). Briefly, protein levels were measured using customized MILLIPLEX MAP® Human Cytokine/Chemokine, Th17, High Sensitivity T Cell, Circulating Cancer Biomarker and Immuno-Oncology Checkpoint Protein Magnetic Bead Panels (Millipore, Billerica, MA) or Human IL-36γ ELISA kit (RayBiotech, Norcross, GA) in accordance with the manufacturer’s protocol. Data were collected with a Bio-Plex® 200 instrument and analyzed using Manager 5.0 software (Bio-Rad, Hercules, CA). The genital inflammatory score system used in this study was described previously (17). Briefly, levels of seven cytokines (IL-1α, IL-1β, IL-8, MIP-1β, MIP-3α, RANTES, and TNFα) were used to determine inflammatory scores; patients were assigned one point for each mediator when the level was in the upper quartile. Patients with inflammatory scores 5-7 were considered to have high genital inflammation, whereas patients with inflammatory scores 1-4 to have low genital inflammation. Patients with inflammatory score 0 were assigned to have no genital inflammation.

### Metabolome analysis

Global metabolome analysis was performed by Metabolon, Inc (Durham, NC) and described previously (29). Briefly, a Waters ACQUITY ultra-performance liquid chromatography (UPLC) and a Thermo Scientific Q-Exactive high resolution/accurate mass spectrometer interfaced with a heated electrospray ionization (HESI-II) source and Orbitrap mass analyzer operated at 35,000 mass resolution were utilized. Metabolites were identified and quantified using Metabolon’s Laboratory Information Management Systems (LIMS).

### Amplicon library preparation and sequencing for microbiome analysis

DNA extraction and 16S rRNA gene sequencing were described previously (17). Briefly, DNA was extracted from vaginal swabs using PowerSoil DNA Isolation Kit (MO BIO Laboratories, Carlsbad, CA) following the manufacturer’s instructions. Amplicon library preparation and sequencing were performed by the Second Genome Inc. (San Francisco, CA). Briefly, the V4 region of bacterial 16S rRNA gene was amplified from the genomic DNA obtained from vaginal swabs and sequenced on the MiSeq platform (Illumina, San Diego, CA).

### Bioinformatics analysis

Microbial DNA sequence data were processed and analyzed using the plugin-based microbiome bioinformatics framework QIIME 2 version 2019.7 (40). DADA2 (41) was used (via the q2-dada2 QIIME 2 plugin) to quality filter the sequence data, removing PhiX, chimeric, and erroneous reads, and merge paired-end reads. Forward and reverse reads were trimmed to 250 nt prior to denoising with dada2, otherwise default parameter settings were used. Taxonomy was assigned to sequence variants using q2-feature-classifier (42) with the classify-sklearn naive Bayes classification method against (1) the GreenGenes 16S rRNA reference database 13_8 release (43) assuming a uniform taxonomic distribution (44); (2) the Genome Taxonomy Database (GTDB) (45), assuming a uniform taxonomic distribution; and (3) GTDB, with taxonomic class weights (expected species distributions) assembled from a collection of 1,017 human cervicovaginal microbiota samples derived from the Vaginal Human Microbiome Project (the same reference set used to construct the STIRRUPS database (46)) using q2-clawback (44). RESCRIPt (https://github.com/bokulich-lab/RESCRIPt) (47) was used to merge these taxonomies via determination of the last common ancestor (LCA) consensus taxonomy assignment for each feature (giving priority to majority classifications, and using superstring matching to facilitate compatibility between the Greengenes and GTDB taxonomies). Any sequence that failed to classify at phylum level was discarded prior to downstream analysis. Microbial feature tables were evenly sampled at 50,000 sequences per sample prior to supervised classification.

Supervised learning was performed in q2-sample-classifier (48) via 10-fold nested cross-validation (classify-samples-ncv method), using random forests classification or regression models [https://doi.org/10.1023/A:1010933404324] grown with 500 trees. Receiver operating characteristic (ROC) curves and area under the curve (AUC) analysis, confusion matrices, and feature importance scores were generated as part of the q2-sample-classifier pipeline. Supervised learning models were trained and tested using the following feature and target data:

1. Disease status was predicted using bacterial 16S rRNA gene ASV abundance, metabolome, and immunoproteome data.
2. *Lactobacillus* dominance was predicted using metabolome and immunoproteome data. *Lactobacillus* dominance categorization was based on the relative frequency of reads classified to genus *Lactobacillus* via 16S rRNA gene sequencing; any sample with ≥ 80% of reads classified as *Lactobacillus* were placed in the *Lactobacillus* dominant (LD) group, and all other samples in the *non-Lactobacillus* dominant (NLD) group.
3. Vaginal pH was predicted using bacterial 16S rRNA gene ASV abundance, metabolome, and immunoproteome data.
4. Genital inflammation scores were predicted using bacterial 16S rRNA gene ASV abundance, metabolome, and immunoproteome data (excluding the 7 immunoproteome markers that are used to calculate the inflammation score).
5. Immunoproteome markers (the abundance of each individual marker) was predicted using metabolome and bacterial 16S rRNA gene ASV abundance data.
6. Metabolite abundance (the abundance of each individual metabolite) was predicted using immunoproteome and bacterial 16S rRNA gene ASV abundance data.

AUC was calculated using scikit-learn (49) for each class, as well as micro- and macro-averages. Micro-average is calculated across each sample, and hence impacted by class imbalances. Macro-average gives equal weight to the classification of each sample, eliminating the impact of class imbalances on average AUC.

Microbe-metabolite interactions were estimated using mmvec (39). This method uses neural networks for estimating microbe-metabolite interactions through their co-occurrence probabilities. Features with fewer than 10 observations were filtered prior to mmvec analysis. Conditional rank probabilities were used to construct principal coordinate analysis biplots (visualized using matplotlib [10.1109/MCSE.2007.55]) that illustrate the co-occurrence probabilities of each metabolite and microbe.

## Results

Interconnection of vaginal microbiome, metabolome, and immune biomarkers Microbe-metabolite interactions were predicted using mmvec (39). This method uses neural networks to estimate microbe-metabolite interactions through their co-occurrence probabilities. This method predicted several strong microbe-metabolite associations. Numerous lipids (including sphingolipids and long-chain unsaturated fatty acids) were associated with multiple amplicon sequence variants (ASVs) belonging to *Prevotella* (including *Prevotella bivia), Peptoniphilus, Streptococcus anginosus, Atopobium vaginae, Sneathia sanguinegenes, Veillonellales, Finegoldia*, and other taxonomic groups (**Figure 2**). *Lactobacillus* ASVs *(Lactobacillus crispatus, Lactobacillus iners, Lactobacillus_H)*, as well as some *Prevotella* (including *Prevotella bivia)*, and other ASVs, were correlated with a range of metabolites including phenylalanylglycine, the anti-inflammatory nucleotide cytosine, glycerophosphoglycerol, glycerol, N-acetyl methionine sulfoxide, and maltopentaose (**Figure 2**). These separations roughly mirror genital inflammation and disease status categories, corresponding with our present findings (described below) as well as previous work showing association between many of these lipids with ICC and high inflammation, and these non-lipid metabolites with high *Lactobacillus* dominance and low inflammation (17, 29). Three-hydroxybutyrate, previously associated with ICC (29), as well as pipecolate, N-acetylcadaverine, and deoxycarnitine were highly correlated with a range of *Streptococcus, Prevotella* (including *Prevotella bivia*), *Megasphaera, Finegoldia, Atopobium vaginae, Sneathia amnii*, and *Sneathia sanguinegens* ASVs. Interestingly, 3-hydroxybutyrate was also correlated to *Lactobacillus iners*.

**Figure 2.**
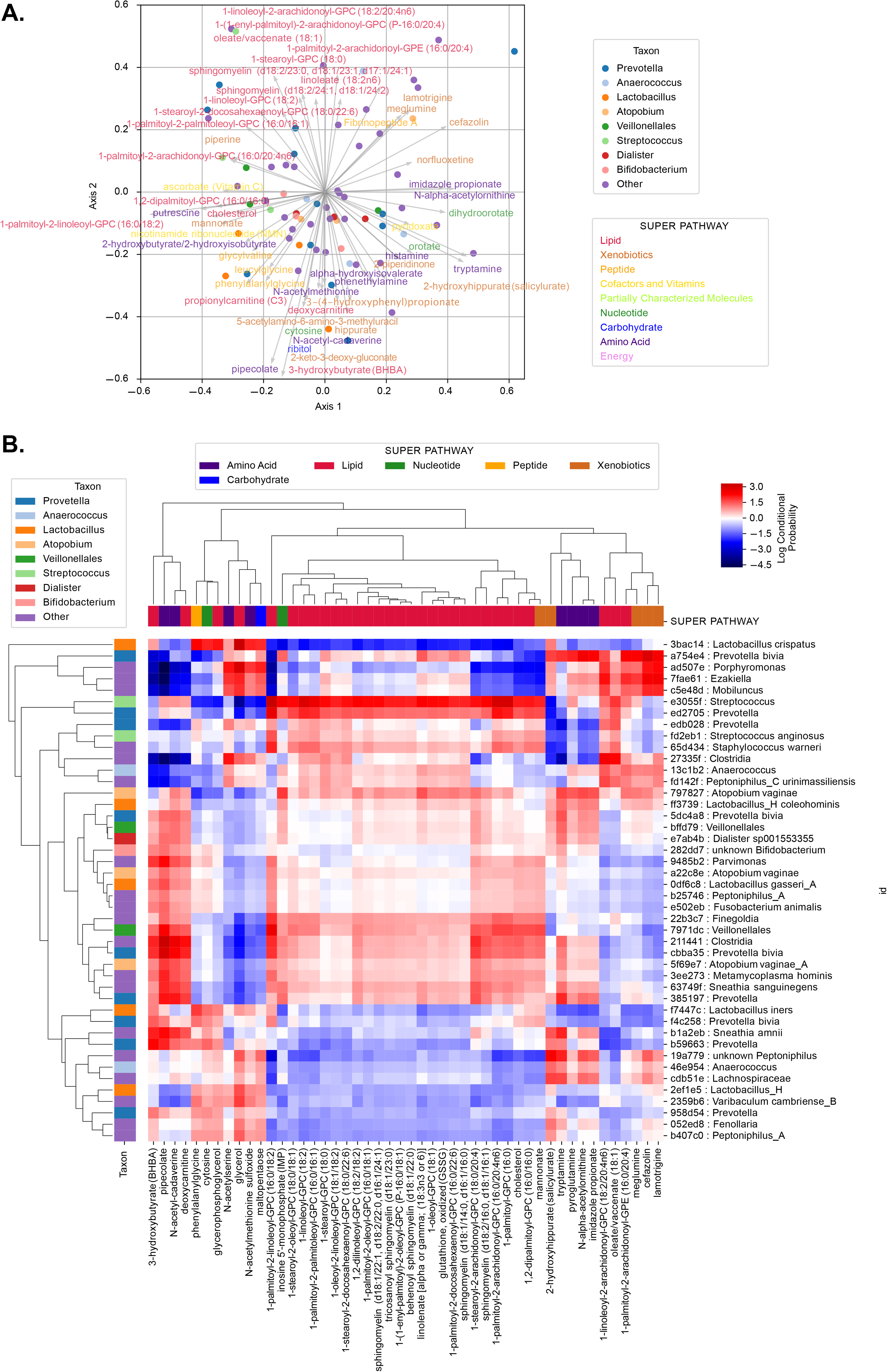
Microbiome-metabolome interaction probabilities via mmvec predicts strong associations between lipid metabolites with *Prevotella, Streptococcus, Atopobium, Sneathia* and other clades [MH1]. **A**. The principal component analysis (PCA) biplot displays the top correlations, colored by genus (for microbial features) or by super pathway (for metabolite features). The correlations were tested using mmvec. This method uses neural networks for estimating microbe-metabolite interactions through their co-occurrence probabilities[MH2]. Microbes (points) and metabolites (arrows) that appear closer to each other in the biplot have a higher likelihood of co-occurring. **B**. The heatmap depicts the correlation coefficients between ASVs and metabolites; hierarchical clustering was done via average weighted Bray-Curtis distance. ASVs were determined using the consensus taxonomy (see Methods section).

To further dissect relationships among the metabolite, microbiome, and immunoproteome, Random Forest regression with 10-fold cross-validation was used to determine the ability to predict the abundance of individual metabolites based on microbiome and immunoproteome profiles, revealing very strong predictive strength for a wide variety of targets (**Supplementary Figure 1, Supplementary Table 1**). This includes the inflammation-and ICC-associated lipids 1-palmitoyl-2-arachidonoyl-gpe (16:0/20:4), 1-palmitoyl-2-linoleoyl-gpc (16:0/18:2), 1,2-dilinoleoyl-gpc (18:2/18:2), 1-palmitoyl-2-docosahexaenoyl-gpc (16:0/22:6), several sphingomyelins, 1-stearoyl-2-docosahexaenoyl-gpc (18:0/22:6), 1-linoleoyl-2-arachidonoyl-gpc (18:2/20:4n6), 1-palmitoyl-2-arachidonoyl-gpc (16:0/20:4n6), arachidonate, and the bile acid glycochenodeoxycholate (**Supplementary Figure 1, Supplementary Table 1**). Many of these associations are driven by high abundances of these lipids, sphingomyelins, and other metabolites in cancer cases: cancer biomarkers are the top predictive features for all of these metabolites (**Supplementary Figure 2**), and when ICC cases are removed from the dataset microbial features (including several *Sneathia, Atopobium, Prevotella, Finegoldia*, and *Mobiluncus* ASVs) are included among the top predictive features, though high predictive strength remains for many (but not all) of these targets (**Supplementary Figure 3-4**). The ability to accurately predict the abundance of these metabolites through cross-validation highlights the close correspondence between the metabolome, microbiome, and immunoproteome across patients, both respective and irrespective of cancer diagnosis.

Random Forest regression was also performed to predict concentration of cancer biomarkers based on microbiome and metabolome profiles, demonstrating strong predictive strength for several targets, including proinflammatory cytokines and chemokines (IL-6, IL-8, IL-36γ, MIF, MIP-1β), the anti-inflammatory cytokine IL-10, growth and angiogenic factors (HGF, SCF, TGF-a,) apoptosis-related proteins (sFAS, TRAIL), the hormone prolactin, the cytokeratin CYFRA21-1, and other cancer biomarkers (AFP, sCD40L, CEA)) (**Supplementary Figure 5**). Metabolites (primarily inflammation-associated lipids) are the most predictive features for each of these targets, but microbial features occur among the top 15 predictive features for many of these, most notably *Adlercreutzia (Eggerthellaceae), Megasphaera, Sneathia*, and *Parvimonas* dominating the top important features for predicting cervicovaginal CEA concentration, regardless of cancer diagnosis (**Supplementary Figure 6**). Several of these biomarkers are clearly related to ICC, as indicated by reduced predictive strength after ICC cases are removed from the dataset; however, most of these markers exhibit similar performance and important feature associations after removing ICC cases (**Supplementary Figures 7-8**).

These findings indicate that both the metabolome and microbiome are highly correlated with and predictive of cancer biomarker concentrations in the cervicovaginal mucosa. Hence, metabolome and microbiome composition can be considered proxy measurements for genital inflammation and immunological responses linked to cervicovaginal carcinogenesis, a relationship that is more explicitly tested below.

### Metabolome and immunoproteome markers predict *Lactobacillus* dominance and vaginal pH

We have previously demonstrated significant negative correlations between *Lactobacillus* dominance (LD), genital inflammation, HPV infection, and ICC (17). Lactobacilli typically dominate the cervicovaginal microbiota of healthy premenopausal women. However, in some women, cervicovaginal microbiota lacks a high proportion of lactobacilli and consists of a consortium of anaerobic bacteria. Intriguingly, Hispanic and black women more frequently exhibit *non-Lactobacillus-dominant* (NLD) microbiota than white or Asian women, which might relate to multiple socioeconomic, environmental and behavioral factors all of which may arise as a result of structural racism (4). LD is associated with low genital inflammation and lower risk of HPV acquisition, persistence and development of precancerous cervical dysplasia (26, 27). Hence, we evaluated the ability of metabolome and immunoproteome features to predict LD, as a proxy for their association with vaginal health in the Arizona-based cohort of women in this study (comprising both non-Hispanic white women (NHW) and women of Hispanic origin). We define LD as any sample in which *Lactobacillus* ASVs collectively comprise ≥ 80% of the vaginal microbiome, and grouped subjects into LD and NLD groups. We then predicted LD status based on metabolome and immunoproteome profile using random forest classification with 10-fold cross-validation. Microbiome data were excluded from the predictive model, as these measurements are non-independent due to compositionality constraints, i.e., changing the relative abundance of one feature (such as a *Lactobacillus* ASV) will alter the relative abundance of other features.

Results demonstrate a very high predictive accuracy (average AUC = 0.94), indicating a near-perfect ability to predict LD or NLD across subjects via cross-validation (**Figure 3A-B**). In other words, cervicovaginal metabolome and immunoproteome profiles are tightly linked to the abundance of *Lactobacillus* spp., suggesting that host immunological response is associated with cervicovaginal microbiome composition. The top predictive features consist primarily of non-lipid metabolites, consistent with the mmvec results (**Figure 2**), though the cancer biomarkers macrophage migration inhibitory factor (MIF) and TNFα also rank among the top 50 most important predictive features (**Figure 3C**). Both MIF and TNFα are more abundant in NLD women (**Supplementary Figure 9**), consistent with higher inflammation and ICC.

**Figure 3.**
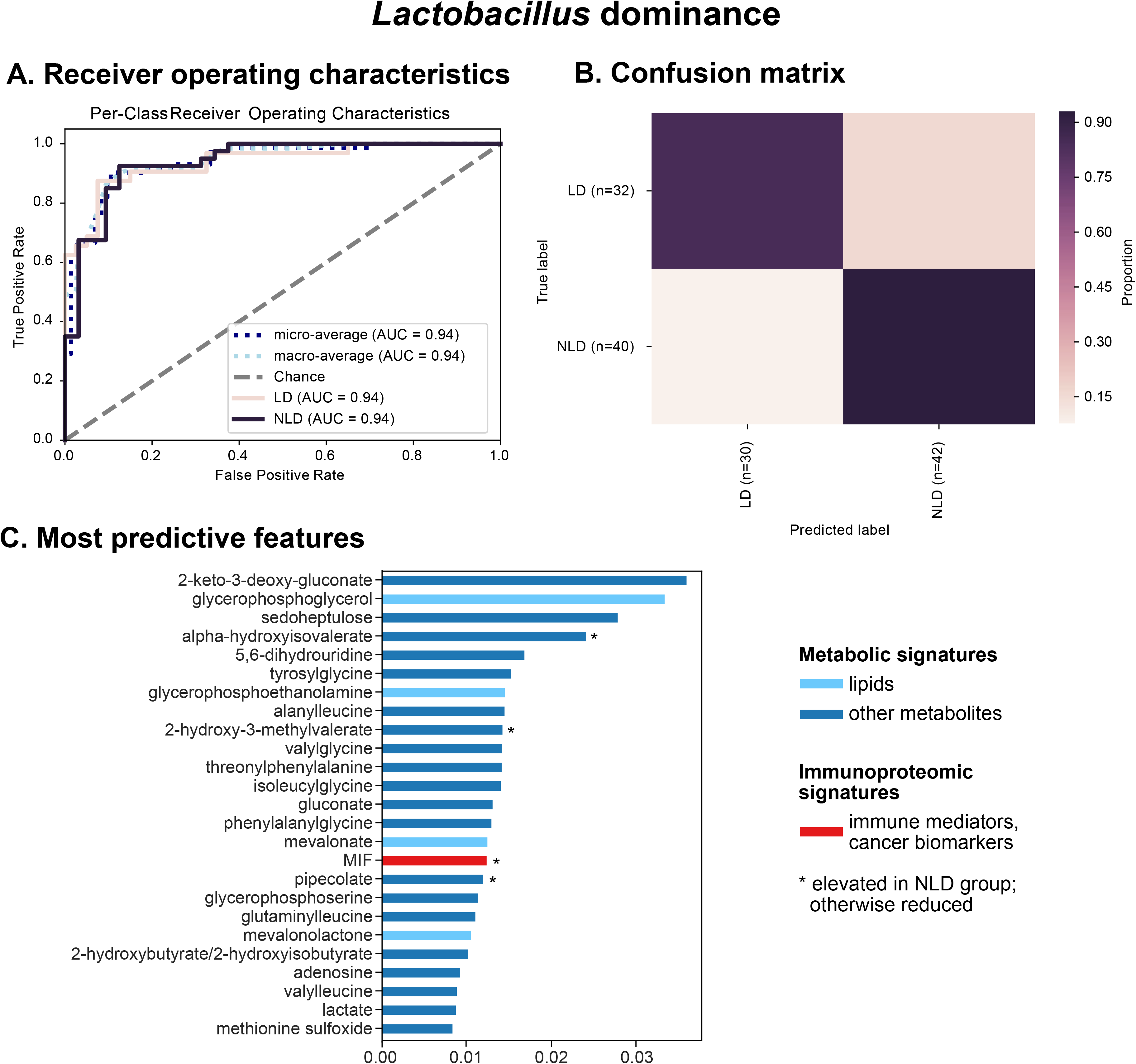
Metabolites (particularly xenobiotics, carbohydrates, amino acids and peptides) and the inflammatory cytokine MIF can accurately predict *Lactobacillus* dominance. Integrated vaginal metabolome and immunoproteome profiles were used as predictive features for training cross-validated Random Forest classifiers to predict whether a subject’s vaginal microbiota is *Lactobacillus* dominant (LD ≥ 80% relative abundance consists of *Lactobacillus* ASVs) or non-LD (NLD < 80% relative abundance consists of lactobacilli). Combined measurements predict the *Lactobacillus* dominance [MH3] at an overall accuracy rate of 88.9%. A 1.6-fold improvement over baseline accuracy was observed. Receiver operating characteristics (ROC) analysis showing true and false positive rates for each group, indicating excellent predictive accuracy for both LD (AUC = 0.94) and NLD groups (AUC = 0.94) (**A**). The confusion matrix illustrates the proportion of times each sample receives the correct classification (**B**). The graphs depict the 25 most strongly predictive features ranked by relative importance score, a measure of their contribution to classifier accuracy (**C**).

Vaginal pH is an important feature of the cervicovaginal microenvironment which relates to *Lactobacillus* dominance. Briefly, vaginal *Lactobacillus* spp. utilize glycogen by-products in the process of fermentation and produce lactic acid, which acidifies the local microenvironment typically to pH below 4.5. This acidic microenvironment contributes to homeostasis and protects the host against invading pathogens and pathobionts. We assessed the predictive relationship between pH and cervicovaginal metabolites, microbiota, and immunoproteome using cross-validated random forest classification models. For the purposes of this analysis, samples were grouped into “low” (pH ≤ 5.0) and “high” pH groups (pH > 5.0). Lower vaginal pH is closely related to demographic characteristics, and Hispanic women tend to have slightly higher average vaginal pH compared to NHW (7, 17), hence we defined pH ≤ 5.0 as “low” for the purposes of this study. Results indicate a weak to moderate predictive relationship (AUC = 070) (**Figure 4A**). Predictive power was lost because a large proportion (35.3%) of women with low vaginal pH were predicted to belong to the high pH group (**Figure 4B**). This characteristic merely indicates that 5.0 is not a reasonable cutoff for the purposes of this analysis; predicting true vaginal pH using a regression model would be more appropriate to characterize the numerical relationship between vaginal pH and the cervicovaginal environment but the small sample size in the current study, strongly skewed toward lower pH values (**Supplementary Figure 10**), prevented the use of cross-validated regression models to evaluate what is likely a more integrative relationship than binary classification can achieve. Results also indicate that this binary pH model, as expected, exhibits many of the same characteristics as the LD/NLD prediction model: many of the same top predictive features were identified (**Figure 4C**). Notably, the top predictive features consist primarily of non-lipid metabolites, and both MIF and TNFα are again in the top 50 most important predictors, both associated with high pH as well as NLD (**Supplementary Figures 9** and **11**). Hence, together these findings recapitulate the associations between LD, low vaginal pH, and low inflammation, and between NLD, high pH, higher inflammation, and carcinogenesis, as well as the microbial and metabolic context of these states, explored in more detail below.

**Figure 4.**
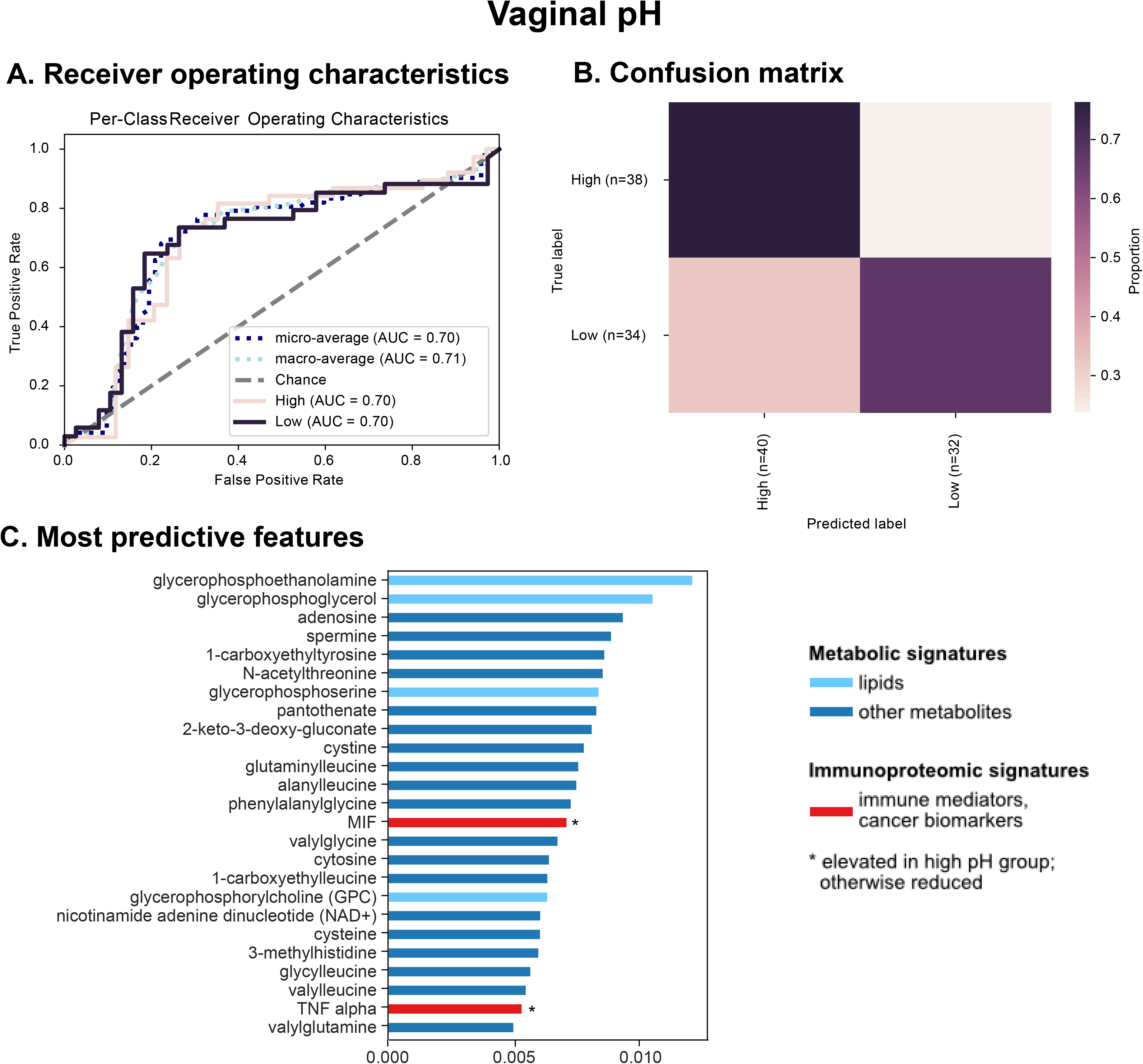
Metabolites (particularly amino acids, peptides and nucleotides) and inflammatory cytokine MIF are the best predictors of vaginal pH. Integrated vaginal microbiome, metabolome, and immunoproteome profiles were used as predictive features for training cross-validated Random Forest classifiers to predict whether a subject’s vaginal pH was low (≤ 5.0) or high (> 5.0). Combined measurements predict vaginal pH at an overall accuracy rate of 72.6%. A 1.4-fold improvement over baseline accuracy was observed. Receiver operating characteristics (ROC) analysis showing true and false positive rates for each group, indicating weak predictive accuracy (micro-average AUC = 0.70) for both low (AUC = 0.70) and high pH groups (AUC = 0.70) (**A**). The confusion matrix illustrates the proportion of times each sample receives the correct classification (**B**). The graphs depict the 25 most strongly predictive features ranked by relative importance score, a measure of their contribution to classifier accuracy (**C**).

### Metabolome, immunoproteome, and microbiome accurately predict genital inflammation but only moderately predict cancer status

Next, we tested the relationship between the cervicovaginal environment and genital inflammation, as a crucial characteristic of ICC progression. We have previously utilized a scoring system to quantify genital inflammation in our cohort (17). To assign genital inflammatory scores (0-7), levels of seven cytokines and chemokines, including IL-1α, IL-1β, IL-8, MIP-1β, MIP-3α, RANTES, and TNFα, were measured in cervicovaginal lavages (CVL) and patients were assigned a score based on whether the level of each immune mediator was in the upper quartile. For the purposes of classification, subjects were grouped into no (score = 0), low (0 < score < 5), or high inflammation (score ≥ 5) groups, and random forest classifiers were trained and tested via 10-fold cross-validation to assess the ability to predict genital inflammation across subjects based on cervicovaginal microbiome, metabolome, and immunoproteome (excluding the 7 inflammatory markers that are used to measure inflammatory score). Results indicate moderately high predictive accuracy (macro-average AUC = 0.86) (**Figure 5A**). Predictive accuracy is very good for high (AUC = 0.93) and no inflammation (AUC = 0.90), but lowest for low inflammation (AUC = 0.75), due to misclassification of some samples as either high or no inflammation (**Figure 5B**). Similar to pH classification but to a lesser extent, this reflects the shortcoming of binning samples for classification into categorical groups, a necessary limitation due to the small sample size of the current study. Regression models predicting actual inflammation score demonstrate high accuracy at lower inflammation scores, but lower accuracy at the upper range due to sparsity of high-inflammation samples for cross-validation (**Supplemental Figure 12**). Larger sample sizes in future studies will enable more accurate prediction of low-inflammation samples through prediction of actual inflammation scores, refining our current estimates of associations between genital inflammation and cervicovaginal microenvironment. As it stands, categorical classification performs moderately well, and can identify a range of features predictive of inflammation, primarily lipids, but also several immune mediators and cancer biomarkers including IL-10, MIP-1α, IL-6, VEGF, and leptin (**Figure 5C, Supplemental Figure 13**).

**Figure 5.**
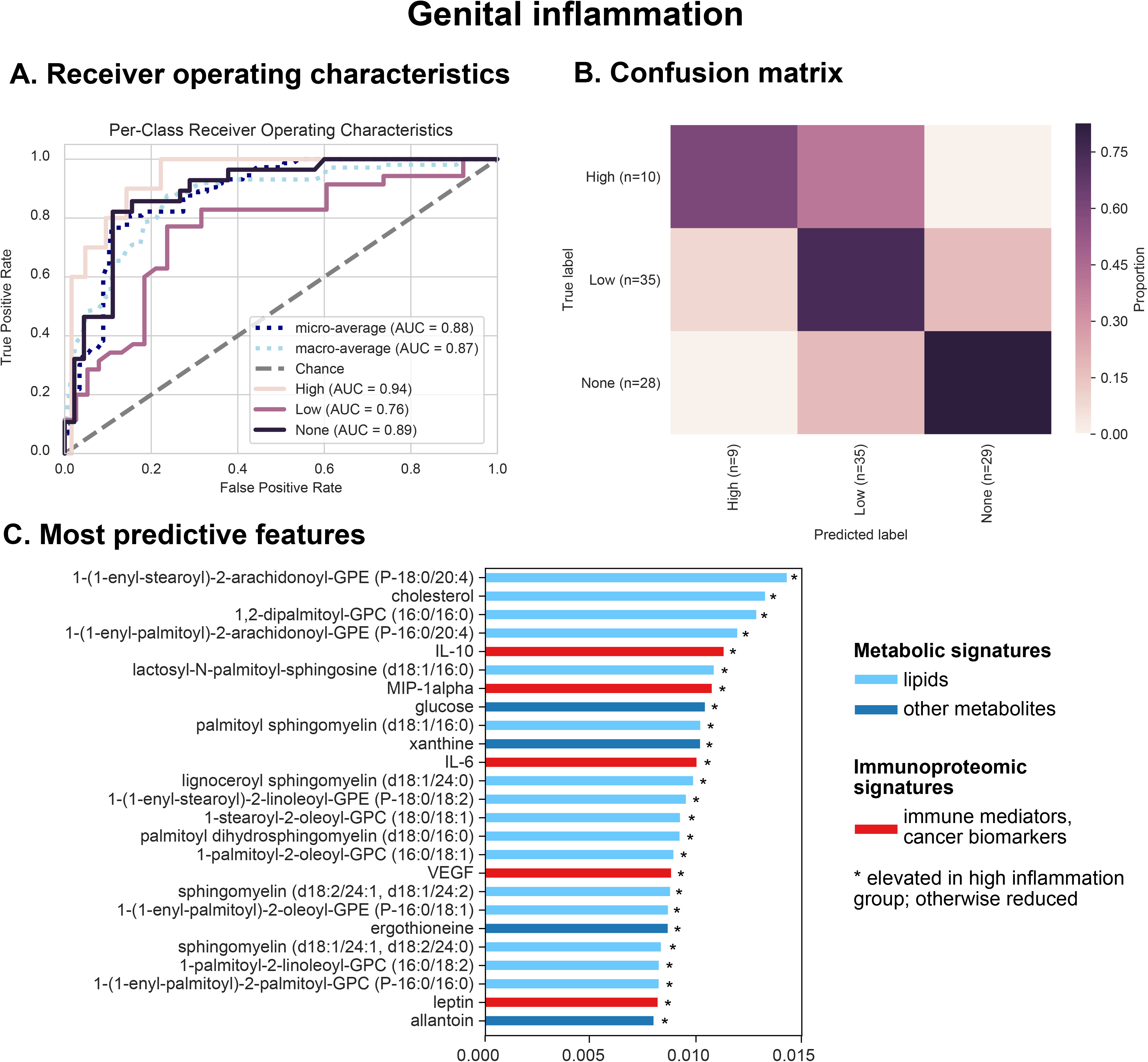
Various metabolites (particularly long-chain fatty acids, sphingolipids and glucose), inflammatory cytokines (IL-6, IL-10, MIP-lalpha) and cancer biomarkers (leptin, VEGF) are the best predictors of the genital inflammation. Integrated vaginal microbiome, metabolome, and immunoproteome profiles (excluding the 7 cytokines used to score genital inflammation) were used as predictive features for training cross-validated Random Forest classifiers to predict whether a subject’s genital inflammation score was “no inflammation” (0), low (1-4), or high (≥ 5.0). Combined measurements predict inflammation score at an overall accuracy rate of 75.3%. A 1.6-fold improvement over baseline accuracy was observed. Receiver operating characteristics (ROC) analysis showing true and false positive rates for each group, indicating moderate average accuracy (micro-average AUC = 0.88) and weak to good predictive accuracy for each group (**A**). The confusion matrix illustrates the proportion of times each sample receives the correct classification (**B**). The graphs depict the 25 most strongly predictive features ranked by relative importance score, a measure of their contribution to classifier accuracy (**C**).

Given the ability to predict genital inflammation, a crucial feature of ICC progression, based on features of the cervicovaginal microenvironment, we sought to determine if HPV infection and carcinogenesis could also be predicted based on these features using cross-validated random forest classification. Samples (*n*=78) were grouped into control HPV- (*n*=18), control HPV+ (*n*=11), LSIL (*n*=12), HSIL (*n*=27), and ICC (*n*=10). This yielded low predictive accuracy (micro-average AUC = 0.73, macro-average AUC = 0.64) (**Supplemental Figure 14**). Although many of the same carcinogenesis-related metabolites and immune markers were top predictors in these models (data not shown), accurate differentiation could not be achieved, primarily because of the low sample size and large class imbalances, but also due to the large number of classes with borderline differences (e.g., high similarity led to misclassification between control HPV– and control HPV+ groups, and between LSIL and HSIL groups). Given the low per-group sample sizes, approaches to mitigate class imbalances were not feasible in the current study, but larger sample sizes and pooled analyses will facilitate better estimates in future studies. However, it should be noted that ICC predictive accuracy was moderately high (AUC = 0.79), in spite of the low sample size and class imbalance (**Supplemental Figure 14**). This indicates that ICC could be predicted with fairly high accuracy across subjects, but non-ICC groups could not be reliably distinguished due to the similarities between these groups. Combining LSIL and HSIL prior to classification increases accuracy, indicating ambiguity between these groups, as reflected in the imprecise distinction between these histological classifications. Hence, ICC elicits signature characteristics in the cervicovaginal microenvironment across subjects that can be used to identify these subjects, but intermediate stages of progression (HPV infection, LSIL, HSIL) cannot be fully distinguished. Larger sample sizes and longitudinal measurement in future studies may improve our ability to diagnose ICC or even predict cancer risk based on cervicovaginal microenvironment characteristics (metabolome, immunoproteome, microbiome).

## Discussion

The vaginal microbiota, HPV infection and cervical neoplasm are related in ways that are still not fully understood. Emerging evidence suggests that *Lactobacillus* dominance (LD) in the vagina and cervix relates to HPV clearance and disease regression, whereas dysbiotic anaerobes contribute to HPV persistence and progression of cervical neoplasm (26-28). Host response to HPV and microbiota, which may result in genital inflammation, immune evasion, and altered metabolism, likely contribute to establishment of persistent infection and disease progression (29, 30, 50-53). Thus, improving our understanding of microbiota-virus-host interactions in the local cervicovaginal microenvironment is imperative for the development of novel diagnostic, preventative and therapeutic approaches, which might help reduce cervical cancer burden among unvaccinated women in the future (54).

We investigated relationships between multiple clinical “omics” datasets (microbiome, vaginal pH, metabolome, immunoproteome) collected from women (who had not been vaccinated against HPV) across cervical carcinogenesis (**Fig. 1**). Using recently developed integrated multi-omics bioinformatics tools, we aimed to establish predictive models and identify key signatures related to vaginal microbiota structure, vaginal pH, genital inflammation and cervical neoplasm status. We identified specific metabolites that were predictive of *Lactobacillus* dominance, vaginal pH, and genital inflammation (**Fig. 3–5**). These findings demonstrate that vaginal microbiota and host defense responses strongly influence cervicovaginal metabolic fingerprints (29, 30, 55) and indicate that cervicovaginal metabolic signatures might be promising biomarkers for gynecological conditions, including cervical cancer. In addition, select immune mediators and cancer biomarkers also exhibited high importance scores in our analyses for predictions of LD and vaginal pH (MIF and TNFα), as well as genital inflammation (IL-6, IL-10, leptin, VEGF), further confirming the link between vaginal microbiota and host immune responses (17, 31, 50, 56, 57). Intriguingly, microbial features did not rank among the top predictors of vaginal pH or genital inflammation. Our neural network analyses and cross-validated Random Forest classification models showed that the abundance of bacterial taxa highly corresponded to levels of key metabolites, immune mediators, and cancer biomarkers related to cervicovaginal health or dysbiosis (**Fig. 2**), suggesting tight coupling of the microbiome, metabolome, and immunoproteome.

Using our approach, we were unable to accurately predict cervical neoplasm status, with the exception of the cervical cancer group, which exhibited a moderate accuracy rate. Relatively low samples size and imbalance in disease classification, which are limitations of our study, might have impacted these predictions. Larger numbers of subjects as well as temporal data on subjects will likely improve predictive models in the future, and better support causal links between microbial dysbiosis and HPV-mediated carcinogenesis. In addition, pathophysiological responses across the continuum of cervical neoplasm might not be uniform among patients with different disease classifications (for example CIN1 and CIN2/3). Indeed, clinical studies have shown contrasting results related to genital inflammation and cervical dysplasia. On one hand, infection with high-risk HPV types or precancerous dysplasia has not been associated with increased level of genital inflammation (17, 50, 53). On the other hand, one report showed increased inflammatory cytokines in patients with cervical dysplasia, but it did not control for microbiota composition (52). Despite not being able to predict disease status, our integrated analyses revealed that we were able to better predict the cervicovaginal microenvironment features.

Our integrated analyses revealed that different classes of metabolites are important for prediction of different phenotypes: lipids were strong predictors of genital inflammation, while amino acids, peptides and nucleotides were predictive of the vaginal microbiota composition. Sphingolipids and long-chain unsaturated fatty acids in particular ranked as top predictors of genital inflammation. Emerging studies have demonstrated that sphingolipids are implicated in multiple pathological processes, such as inflammatory diseases, diabetes, and cancer (58). In a previous report we showed that women with cervical cancer had elevated sphingolipids in the cervicovaginal fluids, suggesting that cancer drives associations of phospholipids with inflammation. However, we observed the correlation with inflammation even after excluding cancer patients (29). In fact, sphingolipids are bioactive metabolites, which may mediate inflammatory signaling through TNFα activation (37). Using neural network analysis, we also showed the co-occurrence of many lipid metabolites and dysbiotic vaginal bacterial taxa (including multiple BV-associated bacteria and *Streptococcus)*, linking microbiota to inflammatory markers.

Predictions of vaginal microbiota and vaginal pH relied mostly on alterations in amino acid metabolism, which was in accordance with previous reports on cervicovaginal metabolomes (30, 36, 55). Specifically we found that 3-hydroxybutyrate (β-hydroxybutyrate, BHB), a ketone body, was strongly correlated with abundance of dysbiotic bacterial species, such as *Streptococcus, Prevotella, Megasphaera, Atopobium* and *Sneathia*, and unexpectedly with one of predominant vaginal *Lactobacillus* spp., *L. iners*. Notably, in a longitudinal clinical study, *L*. iners-dominant vaginal microbiota has been shown to more often transition to dysbiotic NLD microbiota compared to other *Lactobacillus* spp. (59). Furthermore, *L. iners* produces a different ratio of lactic acid isoforms (60), which vary in bactericidal capacities (61); therefore, the protective role of *L. iners* in the cervicovaginal microenvironment is still questionable (62). We have previously demonstrated that 3-hydroxybutyrate (measured in the cervicovaginal fluids) is an excellent discriminator of cervical cancer patients compared to healthy controls (29). Several clinical studies also identified 3-hydroxybutyrate (but measured in serum or tissue effusions) as a potential biomarker of other gynecologic malignancies, such as endometrial cancer (63) and ovarian cancer (64, 65). Three-hydroxybutyrate has also been shown to suppress activation of NLRP3 inflammasome (66). Thus, dysbiotic cervicovaginal bacteria and *L. iners* might utilize this mechanism to evade host defense and, consequently, the inflammasome deregulation might contribute to progression of cervical neoplasm (67).

Other key metabolites that we identify to highly correlate with dysbiotic microbiota were pipecolate and deoxycarnitine. In a previous study on metabolomes of women with BV, these two metabolites positively associated with BV status and the presence of “clue cells” (vaginal squamous epithelial cells covered with bacterial biofilm) (36), which is one of the clinical characteristics of BV. In our report, we also revealed that deoxycarnitine in cervicovaginal fluids can discriminate HPV-positive and HPV-negative women without neoplasia (29), linking vaginal dysbiosis with HPV infection. With regard to the healthy vaginal microbiota, *Lactobacillus* spp. (particularly *L. crispatus)* positively correlated with N-acetyl methionine sulfoxide, a reactive oxygen species. Production of hydrogen peroxide, another reactive oxygen species, by vaginal *Lactobacillus* spp. has been postulated to have a protective effect against invading pathogens (68, 69). Similarly, an increase of N-acetyl methionine sulfoxide in the *Lactobacillus-dominant* cervicovaginal microenvironment might contribute to host protection via oxidative stress.

Through our integrated multi-omics approach, we also identified key immune biomarkers associated with the vaginal microbiota composition and vaginal pH, for instance MIF, a pleiotropic cytokine regulating inflammatory reactions and stress responses (70). MIF was identified as a top predictive factor of vaginal pH and LD in our Random Forest analysis, which took into account multiple different “omics” data types (**Figures 3-4**), suggesting that *Lactobacillus* colonization may be closely involved in regulating markers of genital inflammation, including MIF. In accordance with our finding, several reports have demonstrated significantly increased levels of MIF in cervicovaginal fluids of women with vaginal dysbiosis or BV compared to women with healthy LD microbiota (57, 71, 72). In a previous report we identified MIF (in cervicovaginal fluids) as a potential biomarker discriminating women with cervical cancer from women with dysplasia and healthy controls (31). Other immunohistochemical studies demonstrated overexpression of MIF cervical cancer tissues compared to healthy cervix and dysplasia (73-75). Notably, MIF has been shown to promote cell proliferation, inhibit apoptosis (74) and directly induce secretion of VEGF, an angiogenesis factor (73). Thus, elevated MIF production induced by dysbiotic vaginal microbiota might contribute to cervical carcinogenesis. Our integrated analysis further highlighted the importance of this key immune mediator, and links its expression to vaginal microbiome and metabolome characteristics.

Another pro-inflammatory cytokine that strongly correlated with dysbiotic microbiota and elevated pH was TNFα. Several clinical studies also demonstrated an increase of this cytokine in cervicovaginal fluids of women with vaginal dysbiosis or BV (57, 71, 72, 76). Similar to MIF, microbiota-induced TNFα might enhance cervical carcinogenesis, since this major inflammatory cytokine has been shown to exhibit not only anti-tumor, but also pro-tumor bioactivities (77). Interestingly, *in vitro* studies showed that only particular BV-associated species (for example, *Atopobium vaginae* and *Mobiluncus mulieris*, but not *Prevotella bivia)* induce TNFα production by genital epithelial cells (76, 78-80), suggesting species-specific roles of microbes within dysbiotic polymicrobial consortia on host immunological response, which warrants further investigations. Other immune mediators and cancer biomarkers (IL-6, IL-10, leptin and VEGF) identified to be associated with genital inflammatory scores likely relate to cancer-induced inflammation rather than a host defense response to dysbiotic vaginal microbiota (31). Overall, our data indicate that mucosal inflammation is likely associated with cervical neoplasm via the effect of vaginal microbiota on induction of specific inflammatory mediators and metabolites.

### Integrative omics increases predictive accuracy

Many of the predictive models used in this study integrate multiple omics datasets: metabolome, immunoproteome, and microbiome. We hypothesized that integrating multiple data types would lead to a cumulative increase in predictive accuracy, as accumulating more features could help refine the diagnostic signal of our random forests classifiers, different data types could yield different signature characteristics for the prediction of different subject traits (e.g., inflammation, disease state), and the combined signal could provide more subtle information to differentiate particular groupings of subjects (e.g., LD versus non-LD, disease category). To address this hypothesis directly, we evaluated the performance of each random forest classifier with different combinations of omics data types with the expectation that more data types could only yield better predictive accuracy.

Results indicate that integrating data led to modest increases in accuracy for most classification tasks, but with mixed results (**Figure 6**). For LD, combining multiple datasets led to very modest increases in accuracy (**Figure 6A**). Metabolites alone could predict LD status with high accuracy; immunoproteome data exhibited much poorer accuracy, but combining both data types yielded a slight increase in mean accuracy. For pH prediction, both metabolites and microbiome datasets on their own could predict pH with moderate accuracy, but immunoproteome could not; integrating all three omics datasets led to a slight increase in mean accuracy (**Figure 6B**).

**Figure 6.**
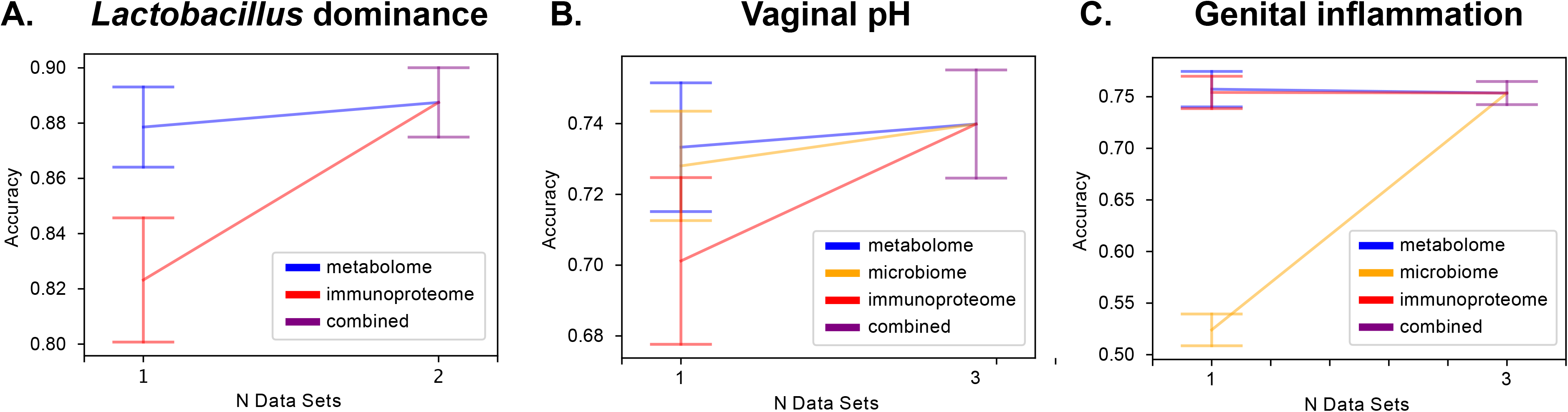
Integrating multiple –omics datasets does not dramatically improve overall prediction accuracy; however, different integration of various measurements are needed for the best prediction of distinct features. Graphs show stepwise accuracy levels for *Lactobacillus* dominance (**A**), vaginal pH (**B**) and genital inflammation (**C**) when random forest models are trained on a single omics dataset or combined data containing 2-3 omics datasets. *Lactobacillus* dominance can be explained mostly by metabolome data, vaginal pH by metabolome and microbiome datasets, and genital inflammation by metabolome and immunoproteome datasets. Combining omics datasets leads to slightly higher average accuracy scores for *Lactobacillus* dominance and vaginal pH classification, but no effect on genital inflammation classification.

Genital inflammation was the one measurement that showed little change in accuracy with integration of multiple omics datasets (**Figure 6C**). Both metabolome and immunoproteome datasets yielded nearly identical high predictive accuracy, whereas microbiome data exhibited poor predictive accuracy. Combining all three datasets led to no change in predictive accuracy. Interestingly, for all tests combining datasets narrowed the variance in accuracy performance (**Figure 6A-C**), suggesting that even if integrating multiple omic data types does not lead to appreciably better accuracy, it could lead to improved reproducibility, but more investigation is required to assess whether this performance enhancement is observed in other studies and disease systems.

### Relevance of a multi-omics approach

Given that we observed only a modest increase in classifier performance accuracy with the use of multiple “omics” data types, it may seem that the benefit of including these additional data does not justify their cost. However, it is important to note that we did not know, *a priori*, which data type would provide the best predictive accuracy in this study. Furthermore, different features types were differentially useful for predicting different features of the cervicovaginal environment. Profiling different feature types therefore enabled discoveries that would not have been possible had we focused only on a single feature type (e.g., the microbiome or the metabolome).

Beginning to collect multi-omics data in human microbiome studies will enable a broader understanding of the complex mechanistic interplay between microbes, metabolites, the host immune system, and host phenotype. We suspect that this additional data will initially improve our ability to make predictions about phenotype, as we have shown in this study. Inspection of our machine learning models to discover important features enables us to develop hypotheses about causation that can be prioritized for evaluation in future studies, and understanding which feature types are most useful in predictive models can provide additional clues for understanding the underlying biology. As our bioinformatics approaches for integrating multi-omics data continue to improve, and as we continue to amass data relating microbes and metabolites to the host immune system and phenotype, we will ultimately improve our ability to model features (such as genital inflammation) based on combinations of microbes and metabolites. This will enable design of treatments based on an understanding of, for example, how the presence of a metabolite will impact the abundance of a group of microbes, which in turn will drive or suppress an immune response.

## Conclusions

There is much work to be done to improve our approaches for integrated multi-omics analyses. For example, developing machine learning classification tools for microbiome multi-omics data that can handle multiple observations per subject to make better use of longitudinal data, and interactive visualization tools that can assist with exploration and interpretation of multi-omics network data will facilitate work. Combining these approaches with novel methods (44) and databases (46, 47) for accurate taxonomic classification of vaginal microbiota will further advance our ability to identify microbial species linked to carcinogenesis and prevention. We posit that integrated multi-omics approaches are essential to enabling many of the advances in human medicine that are promised by microbiome research.

## Data Availability

Bacterial 16s RNA gene sequence data analyzed in this study were deposited in SRA (PRJNA518153). Immunoproteome and metabolome data are available online as supplementary materials accompanying our previous reports (17, 18, 31, 32).

**List of abbreviations**
ASV: amplicon sequencing variants
AUC: area under the curve
BV: bacterial vaginosis
CIN: cervical intraepithelial lesion
Ctrl: control
CVL: cervicovaginal lavage
HPV: human papillomavirus
HSIL: high grade squamous intraepithelial lesions
ICC: invasive cervical carcinoma
LD: *Lactobacillus* dominance
LSIL: low grade squamous intraepithelial lesions
NHW: non-Hispanic white
NLD: *non-Lactobacillus* dominance

## Declarations

### Ethics approval and consent to participate

All participants provided informed written consent and all research and related activities involving human subjects were approved by the Institutional Review Boards at St. Joseph’s Hospital and Medical Center, University of Arizona Cancer Center and Maricopa Integrated Health Systems, all located in Phoenix, AZ.

### Consent for publication

Not applicable.

### Competing interests

Authors declare no competing interests.

### Funding

This study was supported by the Flinn Foundation Grant #1974 to D.M.C. and M.M.H-K., Flinn Foundation Grant #2244 to M.M.H-K. and the National Institutes of Health NCI awards for the Partnership of Native American Cancer Prevention U54CA143924 (UACC) to M.M.H-K and U54CA143925 (NAU) to G.J.C.

### Authors’ contributions

M.M.H.-K. and D.M.C conceived and designed the study. D.M.C. participated in the patient recruitment and sample collection. P.Ł. processed the samples and performed the biological assays. N.A.B. performed bioinformatic analyses. N.A.B., P.Ł., G.J.C. and M.M.H-K. analyzed and interpreted the data. P.Ł and N.A.B. drafted the manuscript. M.M.H-K., G.J.C. and D.M.C. critically reviewed the manuscript. All authors read and approved the final version of the paper.

## Acknowledgements

We would like to thank the patients who enrolled in the study and acknowledge Kelli Williamson, Ann De Jong, Eileen Molzen, Liane Fales, Maureen Sutton for the kind assistance in patient recruitment and sample collection and Drs. Dominique Barnes and Alison Goulder for the assistance with clinical sample and data collection.

## Notes

### Competing Interest Statement

The authors have declared no competing interest.

